# ORAKLE: Optimal Risk prediction for mAke30 in patients with acute Kidney injury using deep Learning

**DOI:** 10.1101/2025.01.18.25320769

**Authors:** Wonsuk Oh, Marinela Veshtaj, Ashwin Sawant, Pulkit Agrawal, Hernando Gomez, Mayte Suarez-Farinas, John Oropello, Roopa Kohli-Seth, Kianoush Kashani, John A. Kellum, Girish Nadkarni, Ankit Sakhuja

## Abstract

**Background:** Major Adverse Kidney Events within 30 days (MAKE30) is an important patient-centered outcome for assessing the impact of acute kidney injury (AKI). The existing prediction models for MAKE30 are static and overlook dynamic changes in clinical status. In this study, we introduce ORAKLE, a novel deep-learning model that utilizes evolving time-series data to predict MAKE30, enabling personalized, patient-centered approaches to AKI management and outcome improvement.

**Methods:** We conducted a retrospective study using three publicly available critical care databases: MIMIC-IV, SICdb, and eICU-CRD. Among these, MIMIC-IV was divided into 80% training and 20% internal test sets, whereas SiCdb and eICU-CRD were used as external validation cohorts. Patients with sepsis-3 criteria who developed AKI within 48 hours of intensive care unit admission were identified. Our primary outcome was MAKE30, defined as a composite of death, new dialysis or persistent kidney dysfunction within 30 days of ICU admission. We developed ORAKLE using Dynamic DeepHit framework for time-series survival analysis and its performance against Cox models using AUROC and AUPRC. We further assessed model calibration using Brier score.

**Results:** We analyzed 16,671 patients from MIMIC-IV, 2,665 from SICdb, and 11,447 from eICU-CRD. ORAKLE outperformed the Cox models in predicting MAKE30, achieving AUROCs of 0.84 (95% CI: 0.83–0.86) vs. in MIMIC-IV internal test set 0.80 (95% CI: 0.78–0.82), 0.83 (95% CI: 0.81–0.85) vs. 0.79 (95% CI: 0.77–0.81) in SICdb, and 0.85 (95% CI: 0.84–0.85) vs. 0.81 (95% CI: 0.80–0.82) in eICU-CRD. The AUPRC values for ORAKLE were also significantly better than that of Cox models. The Brier score for ORAKLE was 0.21 across the internal test set, SICdb, and eICU-CRD, suggesting good calibration.

**Conclusions:** ORAKLE is a robust deep-learning model for predicting MAKE30 in critically ill patients with AKI that utilizes evolving time series data. By incorporating dynamically changing time series features, the model captures the evolving nature of kidney injury, treatment effects, and patient trajectories more accurately. This innovation facilitates tailored risk assessments and identifies varying treatment responses, laying the groundwork for more personalized and effective management approaches.

## INTRODUCTION

Acute kidney injury (AKI) is common among critically ill patients with sepsis. It is associated with high mortality, long-term kidney dysfunction, and a substantial burden on the healthcare system[1–3]. To address the challenges in AKI research and direct the focus towards patient-centered outcomes, the National Institute of Diabetes and Digestive and Kidney Diseases workgroup on Clinical Trials in Acute Kidney Injury recommended a composite outcome measure in 2012[4]. This measure, encompassing death, dialysis initiation, or a sustained increase in serum creatinine, is widely recognized as Major Adverse Kidney Events (MAKE). The MAKE metric provides a valuable benchmark for assessing the impact of AKI on patients.

Despite considerable progress in understanding the pathophysiology of AKI, clinical trials for its management have largely failed to demonstrate efficacy. Advancing both novel and existing treatment strategies requires the ability to accurately identify patients at high risk for meaningful endpoints, such as MAKE. Current management approaches rely on optimizing fluid status, avoiding nephrotoxins, controlling underlying etiologies (e.g., sepsis), and maintaining hemodynamic stability. Our prior work has demonstrated that these approaches can be personalized to better address patient-specific needs by leveraging advanced data-driven methods to refine clinical decision-making[5]. By accurately predicting outcomes like MAKE, we can not only guide personalization of care but also create enriched cohorts for clinical trials, improving the evaluation of both new and existing interventions.

However, current predictive models for MAKE rely on static, predefined time points[6,7], limiting their utility in dynamic and complex clinical environments. These models fail to capture the fluctuating nature of kidney function[8] and critical illness, where patient conditions can evolve rapidly. This study seeks to bridge this gap by introducing ORAKLE (Optimal Risk prediction for mAke30 in patients with acute Kidney injury using deep LEarning), a novel deep learning model to predict the development of MAKE within 30 days (MAKE30). ORAKLE leverages time-series data and event-time modeling to capture the temporal dynamics of patient characteristics and provide robust, individualized risk predictions. By incorporating these advanced techniques, ORAKLE has the potential to revolutionize AKI outcome prediction, enabling dynamic, patient-centered approaches to clinical management and improving outcomes in this high-risk population.

## MATERIALS AND METHODS

### Study Design and Data Sources

We conducted a retrospective observational study utilizing three publicly available critical care databases – Medical Information Mart for Intensive Care (MIMIC-IV) version 3.1, Salzburg Intensive Care Database (SICdb), and eICU Collaborative Research Database (eICU-CRD) version 1.0.8. The MIMIC-IV[9] database comprises electronic health records of critically ill patients admitted to the intensive care units (ICUs) at Beth Israel Deaconess Medical Center (BIDMC) in Boston, MA, USA, between 2008 and 2022. The SICdb[10] includes ICU patient records from University Hospital Salzburg, Austria, spanning 2013 to 2021. The eICU-CRD[11] encompasses de-identified electronic health records of patients admitted to 208 ICUs across the United States during 2014 – 2015. The dataset provides insights into ICU practices and outcomes across diverse healthcare settings.

We divided the MIMIC-IV dataset into two subsets – training set (80%) and internal test set (20%). We utilized SICdb and eICU-CRD databases for independent external validation to evaluate the generalizability of the model.

### Study population

We included adult (age ≥ 18 years), critically ill patients with sepsis, who developed AKI within the first 48 hours of ICU admission. We defined sepsis based on the Third International Consensus Definitions for Sepsis and Septic Shock (Sepsis-3)[12], and AKI based on the Kidney Disease Improving Global Outcomes (KDIGO)[13] guidelines.

We identified patients with sepsis based on a combination of suspected infection, and increase in Sequential Organ Failure Assessment (SOFA)[14] score by ≥ 2 within a 24-hour period[8]. Consistent with prior studies[12,15], we assumed a SOFA score of zero prior to ICU admission. We defined suspicion of infection by concurrent administration of intravenous antibiotics and collection of blood cultures. Specifically, blood cultures must have been collected within 24 hours after intravenous antibiotics were initiated, or intravenous antibiotics must have been administered within 72 hours after the collection of blood cultures[16]. We determined sepsis as the earlier of the suspicion of infection time or SOFA time, provided that the SOFA time occurred no more than 24 hours before or 12 hours after the suspicion of infection[17]. As SICdb and eICU-CRD databases provide limited data on fluid samplings, we relied on the occurrence of antibiotic administrations to define suspicion of infection, a method previously validated in literature[18].

As per KDIGO guidelines, we defined AKI as an increase in serum creatinine of 0.3 mg/dL or more within 48 hours, or a rise of at least 1.5 times the baseline serum creatinine within 7 days[13]. Baseline serum creatinine was determined as the median value measured within 12 months prior to hospital admission. For patients without prior serum creatinine measurements, baseline creatinine was estimated using the Modification of Diet in Renal Disease equation, assuming a glomerular filtration rate of 75 ml/min/1.73 m², as recommended by KDIGO guidelines. Consistent with previous studies, the reference serum creatinine was taken as the lower value between the calculated baseline and the first admission serum creatinine measurement[8,19,20]. We excluded patients with a history of end-stage kidney disease or prior kidney transplantation.

### Outcomes

The primary outcome of interest was major adverse kidney events by 30 days (MAKE30)[21,22], a composite measure defined by 1) death, 2) initiation of new dialysis, or 3) final inpatient serum creatinine ≥ 200% of the reference serum creatinine censored at the earlier of hospital discharge or 30 days after ICU admission.

### Features

We extracted a comprehensive set of predictor variables from the electronic health records, spanning multiple domains of patient information (Table 1 and Supplementary Table S1). These included demographic characteristics (e.g., age, sex, race/ethnicity), vital signs (e.g., heart rate, respiratory rate), laboratory test results (e.g., serum creatinine, blood urea nitrogen [BUN]), severity scores (e.g., AKI stage, Sequential Organ Failure Assessment [SOFA][14] score), and treatment variables (e.g., mechanical ventilation, vasopressor administration).

**Table 1.**
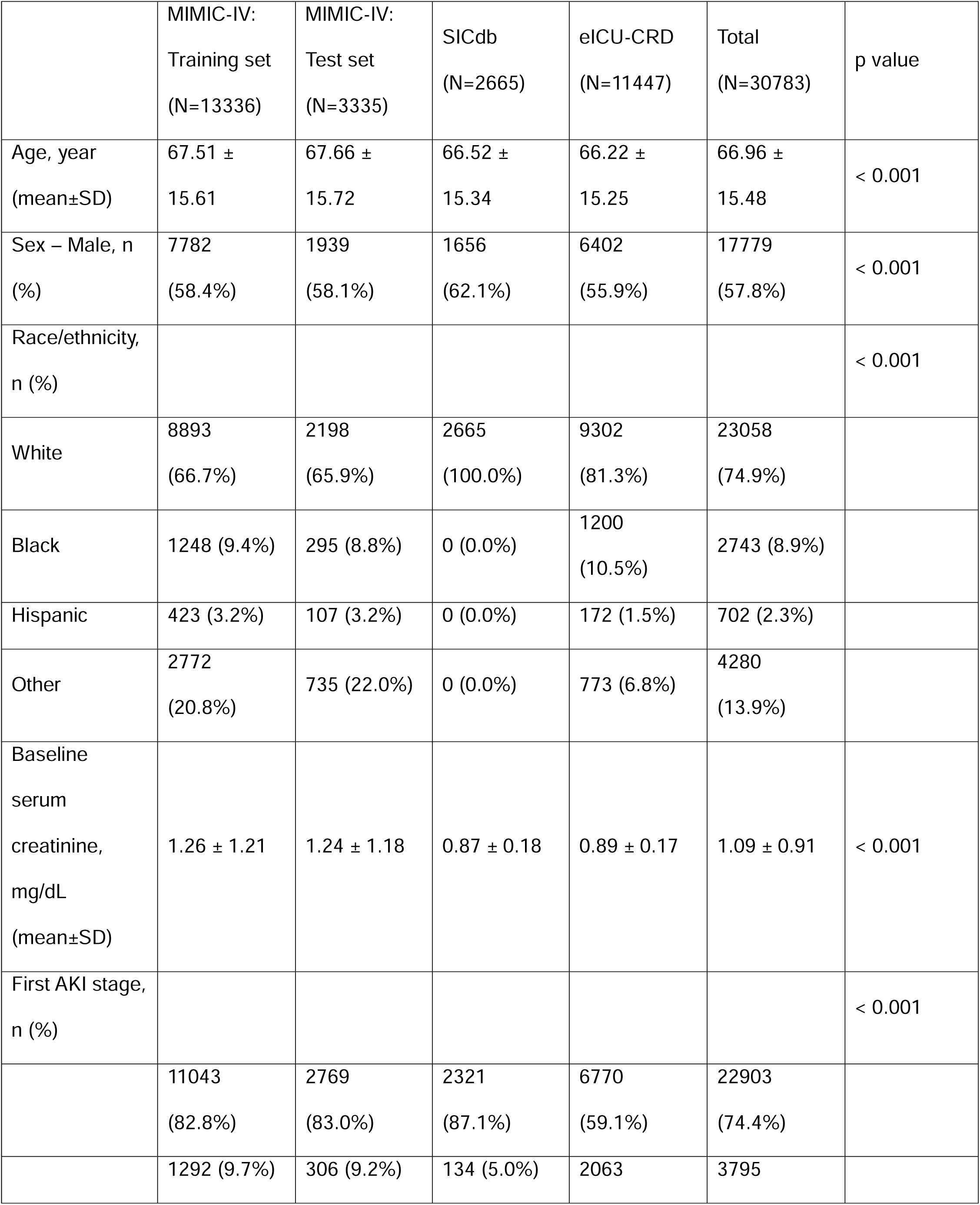
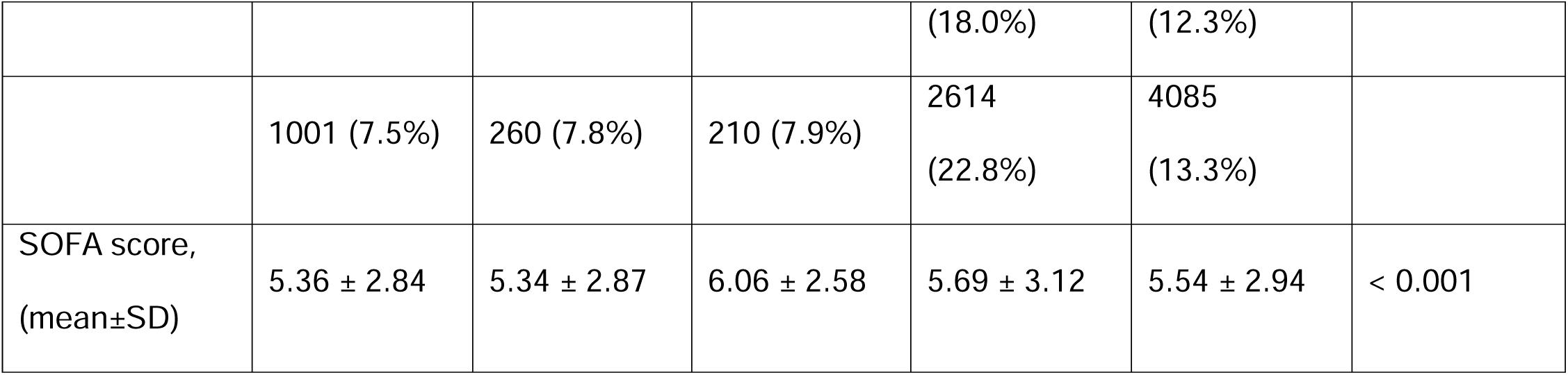
Baseline characteristics of patients in the training and testing sets (MIMIC-IV) and external validation cohorts (SICdb and eICU-CRD).

Only features present in more than 70% of the cohorts were included. Clinical features were summed or averaged within each 8-hour time interval as appropriate. Predictors were constructed for time windows extending from AKI onset (baseline) up to 7 days post-onset. Physiologically implausible outliers were excluded based on clinical expertise. In accordance with standard practices for handling missing data in these datasets, forward fill imputation was applied to all features, except for medications, fluid intake, and urine output which were assigned a value of zero when missing. To address any remaining missing values, we applied multivariate imputation by chained equations (MICE)[23].

### ORAKLE: Model Development and Validation

We used the framework of Dynamic DeepHit[24] to capture the temporal dynamics of patient characteristics. ORAKLE, thus, uses a deep learning survival analysis framework designed to predict time-to-event outcomes while accommodating time-dependent covariates. It has two key subnetworks: 1) a time-series subnetwork, implemented as a Long Short-Term Memory (LSTM) network to model and project multivariate time-series observations effectively; 2) a cause-specific subnetwork, consisting of a fully connected network that outputs the probability of each cause-specific outcome at a given estimation time. The model parameters are trained by maximizing a composite likelihood function, that includes three key components: 1) the likelihood for cause-specific cumulative incidence function, defined as the cumulative probability of observing an event at a specific time divided by the marginal probability of observing an event across all times; 2) the likelihood for ranking cause-specific outcomes, based on the assumption that patients experiencing an event at a particular time should exhibit a higher predicted risk than those who do not; and 3) the likelihood for the time-series model, derived from the product of differences between actual and predicted time-varying covariates. This dual-subnetwork structure enables ORAKLE to capture time-varying patterns in multivariate time-series data and model changes in risk over time simultaneously. Unlike traditional survival models, such as cox regression, which assume proportional hazards or static predictors, ORAKLE captures time-dependent risks dynamically, enabling precise and adaptable risk predictions at any given time point.

We trained ORAKLE using time-series features available at specific time horizons relative to AKI onset. At each time horizon, the time-series features were aggregated and used as input to the model. Predictions were made at 8-hour intervals, starting from AKI onset, and were censored at the earliest of the following events: 7 days post-AKI, transfer out of the ICU, or the development of MAKE30 (Figure 1a).

**Figure 1.**
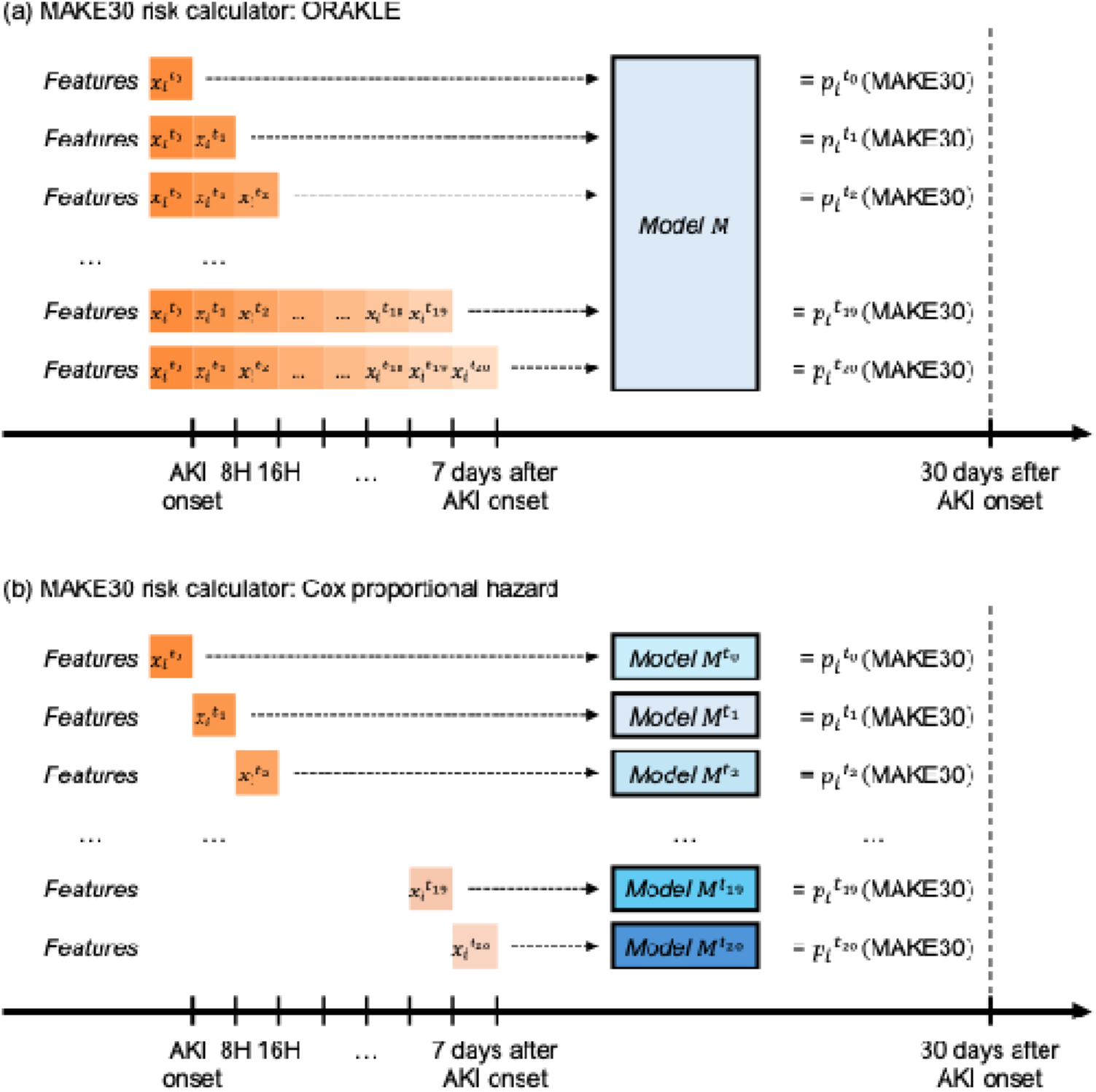
Overview of Study Design. ORAKLE: Optimal Risk prediction for mAke30 in patients with acute Kidney injury using deep LEarning

To evaluate the performance of our MAKE30 prediction model, we compared it against the Cox proportional hazard model as a benchmark. Since the Cox model does not inherently support time-series data, we built separate Cox models for each time horizon. At each specified time point, the models used the available features to estimate the risk of MAKE30 (Figure 1b).

### Statistical analysis

We have reported mean values with standard deviations for continuous features and proportions for categorical variables. We examined the difference across cohorts (MIMIC-IV, SICdb, and eICU-CRD) using analysis of variance (ANOVA) for continuous variables and chi-square tests for categorical variables, with a P value of < .05 considered statistically significant across all statistical tests.

The performance of ORAKLE was compared against the Cox proportional hazards models at each prediction time point. To evaluate predictive performance, we used two key metrics: the area under the receiver operating characteristic curve (AUROC) and the area under the precision-recall curve (AUPRC). AUROC measures the overall discrimination ability by plotting the true positive rate against the false positive rate, offering a comprehensive evaluation of model performance across different thresholds. AUPRC measures the trade-off between precision (the proportion of true positive predictions out of all positive predictions) and recall (the proportion of true positives correctly identified out of all actual positives). We used 10,000 bootstrap resampling to estimate 95% confidence intervals and determine the significance of differences between outcomes.

Additionally, we assessed the calibration performance of the proposed ORAKLE model using the Brier Score, a metric that evaluates the accuracy of probabilistic predictions. To estimate 95% confidence intervals and assess the significance of differences between outcomes, we employed 10,000 bootstrap resampling iterations.

To allow for explainability of predictions of the model, we assessed for feature importance using SHapley Additive exPlanations (SHAP) values. Mean absolute SHAP values quantify the influence of individual predictors on the outcome (MAKE30), with higher values indicating greater importance in model predictions and lower values reflecting lesser influence.

All statistical analyses were performed using Python, version 3.6.13, with TensorFlow, version 1.13, and R software, version 4.3.0.

## RESULTS

### Study population

This study employed three distinct databases to develop and validate, ORAKLE, a deep learning model to predict major adverse kidney events by 30 days (MAKE30) in a continuous fashion. From the MIMIC-IV database, 16,671 patients satisfied the inclusion criteria, with 13,336 (80) used for model development and 3,335 (20) as internal test cohort (Supplemental Figure S1). External validation was conducted using two independent cohorts: 2,665 patients from the SICdb database and 11,447 patients from the eICU-CRD database. The mean age of patients in the MIMIC-IV cohort was 67.5 years, with 58.3 male. The SICdb cohort had a mean age of 66.5 years, with 62.1 males. In comparison, the eICU-CRD cohort had a mean age of 66.2 years, with 55.9 males. A detailed summary of baseline characteristics for all cohorts is presented in Table 1 and Supplementary Table S1. The overall incidence of MAKE30 was 29 in MIMIC-IV, 26 in SiCdb and 39 in eICU-CRD.

### Performance of ORAKLE

ORAKLE consistently outperformed the Cox proportional hazard model in prediction of MAKE30 (Figure 2). For the internal test set, ORAKLE achieved an overall AUROC of 0.84 (95% CI: 0.83–0.86), significantly higher than the Cox model’s macro-averaged AUROC of 0.80 (95% CI: 0.78–0.82) with a p-value < 0.001. This performance advantage was consistent across external validation cohorts. In the SICdb cohort, ORAKLE achieved an AUROC of 0.83 (95% CI: 0.81–0.85) compared to 0.79 (95% CI: 0.77–0.81) for the Cox model, while in the eICU-CRD cohort, ORAKLE achieved an AUROC of 0.85 (95% CI: 0.84–0.85) compared to 0.81 (95% CI: 0.80–0.82). All differences were statistically significant, with p-values < 0.001.

**Figure 2.**
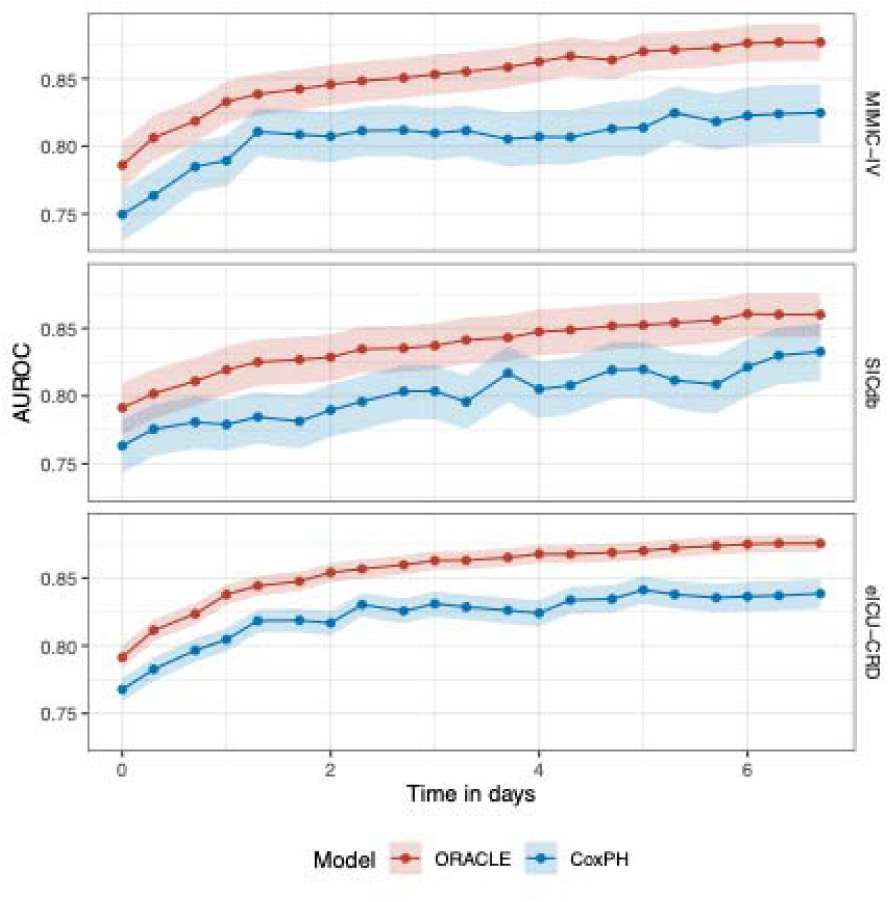
Comparative AUROC for ORAKLE and Cox proportional hazard models in predicting MAKE30 across time horizons.

At the onset of AKI, the ORAKLE demonstrated clear advantages in predictive performance compared to the Cox proportional hazard model. In the internal test set, ORAKLE achieved an AUROC of 0.79 (95% CI: 0.77–0.80), significantly outperforming the Cox model’s AUROC of 0.75 (95% CI: 0.73–0.77), with a p-value < 0.001. This trend persisted in external validation cohorts, where ORAKLE achieved an AUROC of 0.79 (95% CI: 0.77–0.81) in the SICdb cohort compared to 0.76 (95% CI: 0.74–0.78) for the Cox model, and an AUROC of 0.79 (95% CI: 0.78–0.80) in eICU-CRD cohort compared to 0.77 (95% CI: 0.76–0.78) for the Cox model, with p-values < 0.001 across all comparisons.

The model performance steadily improved predictive performance as the prediction time point moved further from the onset of AKI. Both ORAKLE and the Cox proportional hazards model showed increasing AUROC values as the prediction window progressed from AKI onset to 7 days post-onset across both the internal test set and external validation cohorts. However, the performance gap between the two models widened over time, highlighting ORAKLE’s superior ability to utilize extended time-series data for more accurate and dynamic risk prediction.

ORAKLE also consistently outperformed the Cox proportional hazards model in terms of AUPRC. The overall AUPRC for ORAKLE was 0.74 (95% CI: 0.71–0.76), 0.71 (95% CI: 0.68–0.74), and 0.80 (95% CI: 0.79–0.81) in the MIMIC-IV, SICdb, and eICU-CRD cohorts, respectively, compared to 0.64 (95% CI: 0.60–0.68), 0.58 (95% CI: 0.54–0.62), and 0.72 (95% CI: 0.71–0.74) for the Cox model. At the onset of AKI, the AUPRC for ORAKLE was 0.61 (95% CI: 0.58–0.65) in the internal test set, compared to 0.58 (95% CI: 0.54–0.61) for the Cox model (p < 0.001). While both models demonstrated improved performance as the prediction time point moved further from AKI onset, ORAKLE exhibited more substantial gains, reflecting its enhanced ability to utilize time-series data. By day 7, the ORAKLE achieved an AUPRC of 0.79 (95% CI: 0.77–0.81) versus 0.68 (95% CI: 0.64–0.72) for the Cox model in internal test set, with p-values < 0.001 for all comparisons (Figure 3).

**Figure 3.**
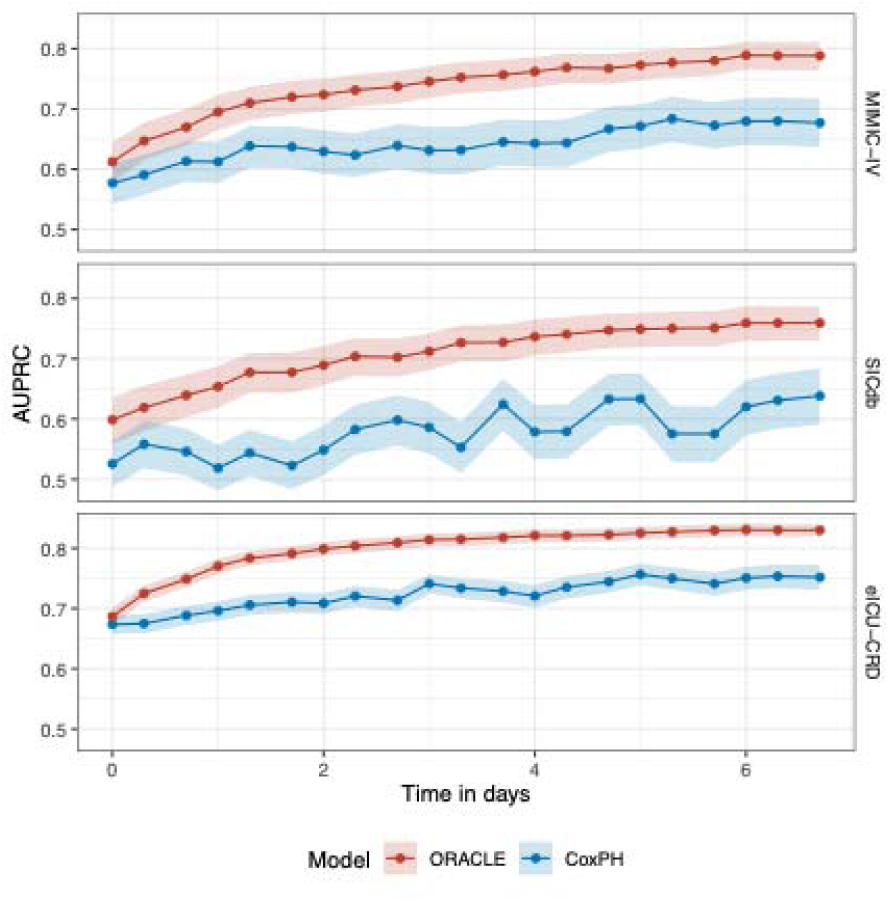
Comparative AUPRC for ORAKLE and Cox proportional hazard models in predicting MAKE30 across time horizons.

Similar trends were observed in external cohorts. At AKI onset, ORAKLE’s AUPRC was 0.60 (95% CI: 0.56–0.64) in the SICdb cohort and 0.69 (95% CI: 0.67–0.70) in the eICU-CRD cohort, compared to 0.53 (95% CI: 0.49–0.56) and 0.67 (95% CI: 0.66–0.69) for the Cox model, respectively. By day 7, ORAKLE achieved an AUPRC of 0.76 (95% CI: 0.73–0.79) in the SICdb cohort and 0.83 (95% CI: 0.82–0.84) in the eICU-CRD cohort, outperforming the Cox model’s AUPRC of 0.64 (95% CI: 0.59–0.68) and 0.75 (95% CI: 0.73–0.77), respectively.

Finally, we evaluated the calibration of the ORAKLE using the Brier Score. The Brier score for ORAKLE was 0.21 (95% CI: 0.21 – 0.21) in internal test set, 0.21 (95% CI: 0.20 – 0.21) in SICdb and 0.21 (95% CI: 0.20 – 0.21) in eICU-CRD. Figure 4a shows the model’s Calibration Plot and Figure 4b illustrates the Brier Scores for ORAKLE across three cohorts over time.

### Feature Importance

At the time of prediction, AKI stage emerged as the most influential variable for predicting MAKE30, followed by age, patient weight, serum creatinine, and the SOFA score (Figure 4).

**Figure 4.**
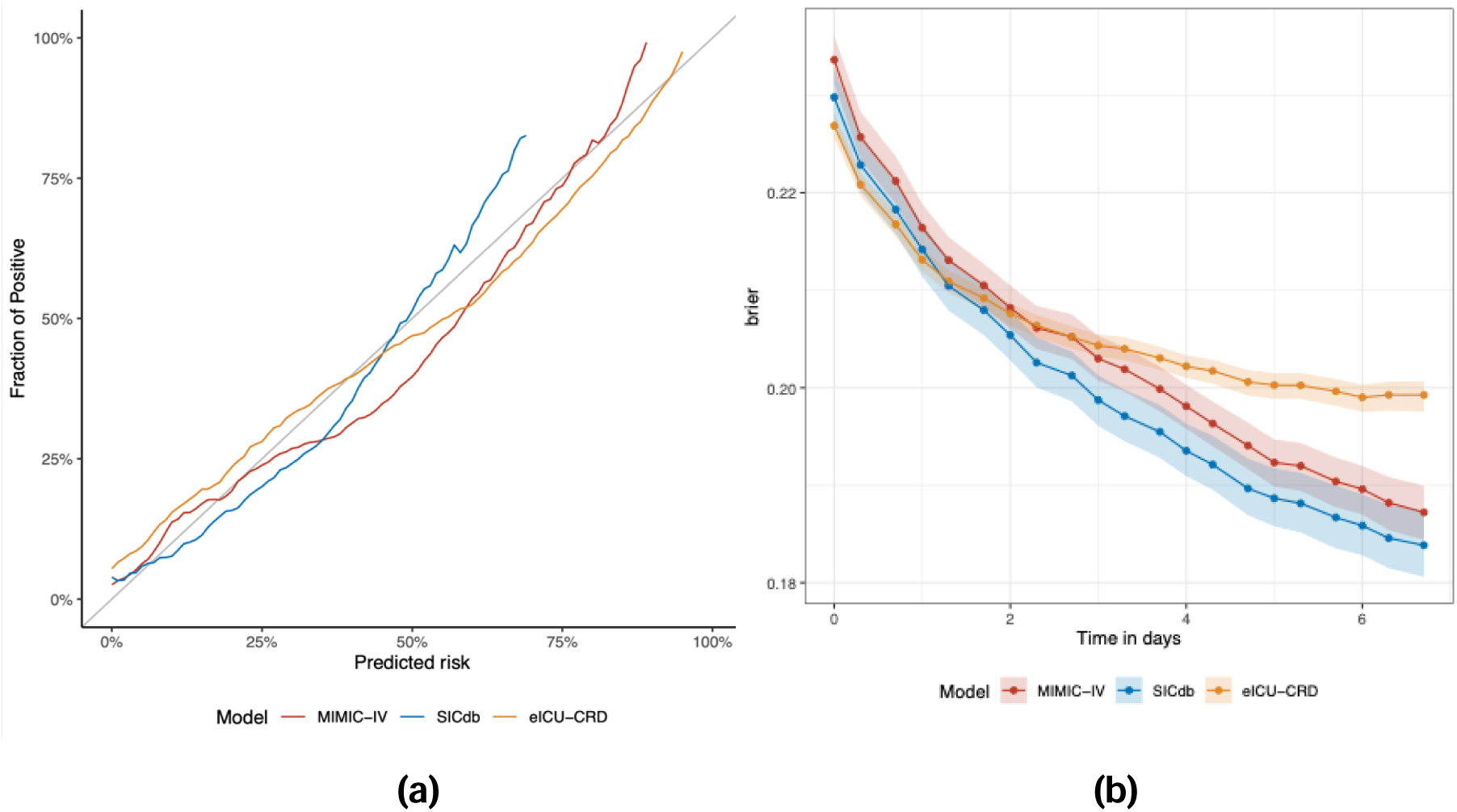
ORAKLE’s a) Calibration Plot, b) Brier Scores over time.

**Figure 5.**
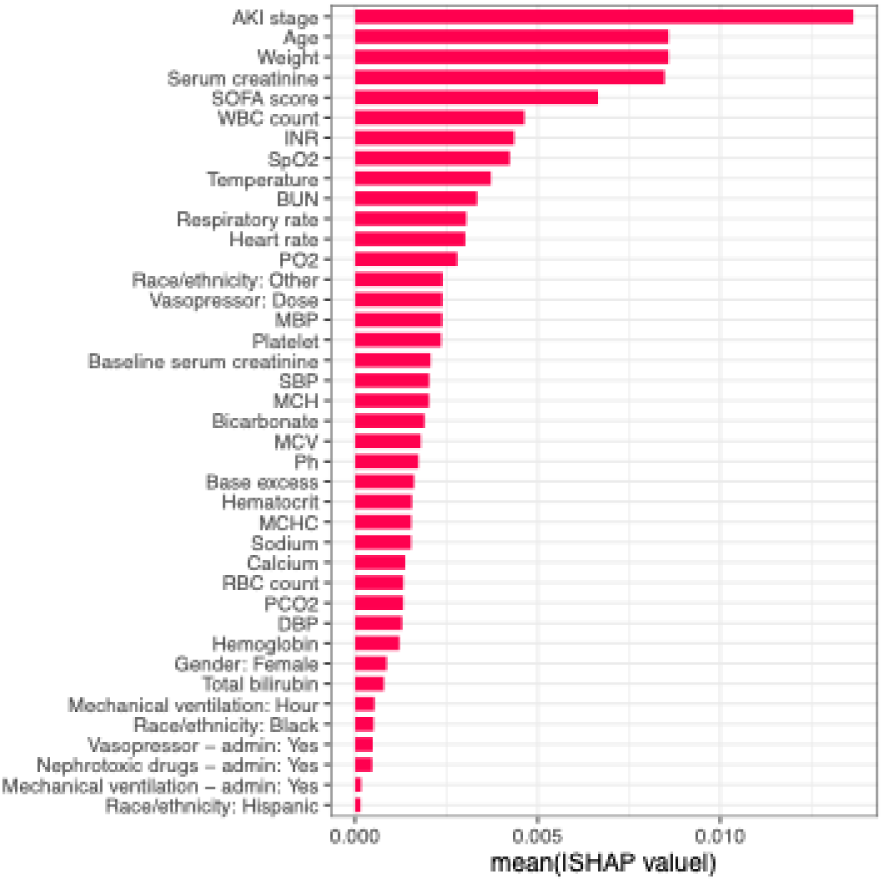
Feature importance for MAKE30 in the ORAKLE. Features: Age, Race/ethnicity: Black, Race/ethnicity: Hispanic, Race/ethnicity: Other, Gender: Female, Heart rate, SBP, DBP, MBP, Respiratory rate, Temperature, SpO2, Weight, Hematocrit, Hemoglobin, MCH, MCHC, MCV, Platelet, RBC count, WBC count, Bicarbonate, BUN, Calcium, Serum creatinine, Sodium, INR, Total bilirubin, PO2, PCO2, Ph, Base excess, Baseline serum creatinine, AKI stage, SOFA score, Vasopressor – admin: Yes, Vasopressor: Dose, Mechanical ventilation – admin: Yes, Mechanical ventilation: Hour, Nephrotoxic drugs – admin: Yes

## DISCUSSION

In this study, we developed ORAKLE, a deep learning model designed for dynamic prediction of MAKE30, leveraging the evolving clinical characteristics of patients over time. The model generates predictions every 8 hours, starting at AKI onset and continuing up to the first 7 days post-AKI. This 8-hour interval balances the need for timely updates with sufficient time for clinicians to make informed decisions, ensuring clinical relevance. ORAKLE demonstrated excellent performance, with high AUROC and AUPRC values, and has been validated across diverse datasets – a heterogeneous U.S. cohort and a European ICU cohort. This robust validation underscores the model’s generalizability and potential to support dynamic, actionable decision-making in varied clinical settings.

Most predictive models in the field of AKI have focused primarily on predicting the development of AKI itself[25]. However, among patients who already have AKI, it is crucial to identify those at high risk for adverse outcomes such as MAKE. Understanding this risk is vital for tailoring existing therapies to individual patient needs and for selecting enriched patient samples for clinical trials of new or existing interventions. The KDIGO guidelines provide a framework for AKI management, and their implementation has been shown to improve outcomes in randomized controlled trial setting[26]. However, compliance with these guidelines is notably low in routine clinical practice[27,28], largely due to their nonspecific nature and the challenge of determining which patients would benefit most from specific components of the recommendations. Our prior work has demonstrated that specific consensus-based recommendations for management of AKI can be developed for subgroup of patients[29], demonstrating that this limitation in guidelines can be addressed. Nonetheless, the critical next step lies in identifying the differential effects of treatments for individual patients. Achieving this requires dynamic, time-sensitive risk assessment for adverse outcomes such as MAKE30. By enabling this evolving risk assessment, we can not only refine personalized treatment strategies but also optimize the evaluation of therapies, paving the way for truly individualized AKI management.

There is a paucity of studies focused on predicting MAKE, and none of the available models incorporate evolving time-series data into their predictions. For instance, Neyra and colleagues compared the performance of various machine learning algorithms such as logistic regression, random forest, support vector machine, and extreme gradient boosting to predict MAKE at 120 days[7]. While their model demonstrated good performance with an AUC of 0.73 in an external validation cohort, it did not account for time-varying covariates and was limited to critically ill adult patients during the first three days of hospitalization, restricting its generalizability. Similarly, Xiaoyuan and colleagues developed an early warning model for MAKE30 using a nomogram based on variables collected within the first 24 hours of ICU admission[30]. Their model exhibited good predictive performance, with AUC of 0.82 in external validation cohort but was limited to predictions within first day of ICU admission and did not incorporate temporally evolving data. This is particularly critical in the ICU, where a patient’s clinical condition is highly dynamic. Our prior work has demonstrated that changes in creatinine over time are a significant predictor of both acute kidney disease and mortality[8]. Therefore, capturing this dynamic evolution in clinical characteristics is essential for accurately predicting outcomes in AKI patients. Unlike static models that rely on fixed data points captured at specific intervals, ORAKLE employs a double subnetwork architecture designed for dynamic prediction. It utilizes an LSTM network to process multivariate time-series data, coupled with a fully connected cause-specific network that estimates probabilities for each outcome at a given time. This innovative architecture provides a deeper and more nuanced understanding of AKI progression, effectively addressing the limitations inherent in static prediction models.

LSTMs, are a type of recurrent neural network (RNN) designed to handle time-series data by addressing the vanishing and exploding gradient problems of traditional RNNs[31,32]. Unlike feedforward networks, which process inputs independently, RNNs maintain “memory” through hidden states, enabling them to model temporal and sequential dependencies. However, standard RNNs often struggle to retain long-term information. LSTMs resolve these limitations through their unique architecture, which includes memory cells and gating mechanisms. These components enable the selective retention, updating, and removal of information, allowing LSTMs to maintain relevant context over extended sequences. This capability makes them particularly effective for tasks like time-series prediction. In clinical applications, LSTMs excel at modeling time-dependent health data such as electronic health records (EHRs) and continuous monitoring outputs, integrating both abstract and temporally distant variables[33]. Unlike traditional models like Cox regression, which struggle to model dynamic relationships over time, LSTMs are specifically designed to handle sequential data with changing patterns. This is especially true when the changes are nonlinear or when there are intricate, time-dependent dependencies that are not well captured by the Cox model. This makes LSTMs a superior choice for modeling outcomes in diseases like AKI, where kidney function can vary rapidly and treatment decisions need to be adjusted continuously based on the evolving patient data.

While ORAKLE demonstrates strong performance, it is important to acknowledge its limitations. First, as a retrospective analysis, it is inherently susceptible to biases, such as selection bias and potential confounding factors, which could influence the interpretation of the results. However, the study demonstrated good generalizability, as evidenced by its strong performance in both internal and two distinct external validation datasets. Second, while the model shows strong performance, it has yet to be deployed prospectively in real-world clinical environments. While the model provides an easily deployable framework, the next step is to implement it in clinical practice to evaluate its effectiveness in a dynamic, clinical context. Third, our sample consists of patients who developed AKI within the first 48 hours of ICU admission, which may limit the generalizability of the findings to all patients with AKI or those who develop AKI outside of the ICU setting. Finally, our AKI definition did not include patients with isolated oliguria, and as a result, the findings may not be applicable to this subgroup of patients.

In conclusion, we have developed a robust model that marks a significant advancement in MAKE30 prediction and AKI management. By integrating dynamically evolving time-series features, the model offers a more accurate reflection of the dynamic nature of kidney injury, treatment responses, and patient progression. This approach enables individualized risk assessments and the identification of differential responses to treatments, paving the way for more personalized and effective management strategies.

## DECLARATIONS

### Ethics approval and consent to participate

The study received approval from the Institutional Review Board at the Icahn School of Medicine at Mount Sinai (approval no. 19-00951).

### Availability of data and materials

Publicly available datasets were analyzed in this study. The MIMIC-IV dataset is available at https://physionet.org/content/mimiciv/, the eICU-CRD dataset is available at https://physionet.org/content/eicu-crd/, and the SICdb dataset is available at https://physionet.org/content/sicdb/.

### Authors’ contributions

WO and AS designed and conceptualized the study. WO and AS performed data collection and analysis. WO, MV, and AS wrote the first draft of the manuscript. All authors interpreted the data analyses, critically revised the manuscript for important intellectual content, and read and approved the final version. All authors gave final approval for the version to be published. WO is first author, GNN and AS provided supervision for the study as co-senior authors.

## ABBREVIATIONS

MAKE: Major Adverse Kidney Events
MAKE30: Major Adverse Kidney Events within 30 days
AKI: Acute Kidney Injury
ORAKLE: Optimal Risk prediction for mAke30 in patients with acute Kidney injury using deep Learning
MIMIC-IV: Medical Information Mart for Intensive Care
SICdb: Salzburg Intensive Care Database
eICU-CRD: eICU Collaborative Research Database
ICU: Intensive Care Unit
BIDMC: Beth Israel Deaconess Medical Center
SOFA: Sequential Organ Failure Assessment
Sepsis-3: Third International Consensus Definitions for Sepsis and Septic Shock
MICE: Multivariate Imputation by Chained Equations
LSTM: Long Short-Term Memory
ANOVA: ANalysis Of Variance
AUROC: Area Under the Receiver Operating Characteristic curve
AUPRC: Area Under the Precision-Recall Curve
SHAP: SHapley Additive exPlanations
EHR: Electronic Health Records
RNN: Recurrent Neural Network
KDIGO: Kidney Disease: Improving Global Outcomes

## REFERENCES

1. Coca SG, Singanamala S, Parikh CR. Chronic kidney disease after acute kidney injury: a systematic review and meta-analysis. Kidney International. 2012;81:442–8.

2. Hoste EAJ, Bagshaw SM, Bellomo R, Cely CM, Colman R, Cruz DN, et al. Epidemiology of acute kidney injury in critically ill patients: the multinational AKI-EPI study. Intensive Care Med. 2015;41:1411–23.

3. Dasta JF, Kane-Gill SL, Durtschi AJ, Pathak DS, Kellum JA. Costs and outcomes of acute kidney injury (AKI) following cardiac surgery. Nephrology Dialysis Transplantation. 2008;23:1970–4.

4. Palevsky PM, Molitoris BA, Okusa MD, Levin A, Waikar SS, Wald R, et al. Design of Clinical Trials in Acute Kidney Injury: Report from an NIDDK Workshop on Trial Methodology. Clinical Journal of the American Society of Nephrology. 2012;7:844–50.

5. Oh W, Takkavatakarn K, Kittrell H, Shawwa K, Gomez H, Sawant AS, et al. Development and Validation of a Policy Tree Approach for Optimizing Intravenous Fluids in Critically Ill Patients with Sepsis and Acute Kidney Injury [Internet]. Intensive Care and Critical Care Medicine; 2024 [cited 2025 Jan 2]. Available from: http://medrxiv.org/lookup/doi/10.1101/2024.08.06.24311556

6. McKown AC, Wang L, Wanderer JP, Ehrenfeld J, Rice TW, Bernard GR, et al. Predicting Major Adverse Kidney Events among Critically Ill Adults Using the Electronic Health Record. J Med Syst. 2017;41:156.

7. Neyra JA, Ortiz-Soriano V, Liu LJ, Smith TD, Li X, Xie D, et al. Prediction of Mortality and Major Adverse Kidney Events in Critically Ill Patients With Acute Kidney Injury. American Journal of Kidney Diseases. 2023;81:36–47.

8. Takkavatakarn K, Oh W, Chan L, Hofer I, Shawwa K, Kraft M, et al. Machine learning derived serum creatinine trajectories in acute kidney injury in critically ill patients with sepsis. Crit Care. 2024;28:156.

9. Johnson AEW, Bulgarelli L, Shen L, Gayles A, Shammout A, Horng S, et al. MIMIC-IV, a freely accessible electronic health record dataset. Sci Data. 2023;10:1.

10. Rodemund N, Wernly B, Jung C, Cozowicz C, Koköfer A. Harnessing Big Data in Critical Care: Exploring a new European Dataset. Sci Data. 2024;11:320.

11. Pollard TJ, Johnson AEW, Raffa JD, Celi LA, Mark RG, Badawi O. The eICU Collaborative Research Database, a freely available multi-center database for critical care research. Sci Data. 2018;5:180178.

12. Singer M, Deutschman CS, Seymour CW, Shankar-Hari M, Annane D, Bauer M, et al. The Third International Consensus Definitions for Sepsis and Septic Shock (Sepsis-3). JAMA. 2016;315:801.

13. Khwaja A. KDIGO Clinical Practice Guideline for Acute Kidney Injury. Kidney International Supplements. 2012;2:1–138.

14. Vincent J, Moreno R, Takala J, Willatts S, De Mendonça A, Bruining H, et al. The SOFA (Sepsis-related Organ Failure Assessment) score to describe organ dysfunction/failure. Intensive Care Medicine. 1996;22:707–10.

15. Raith EP, Udy AA, Bailey M, McGloughlin S, MacIsaac C, Bellomo R, et al. Prognostic Accuracy of the SOFA Score, SIRS Criteria, and qSOFA Score for In-Hospital Mortality Among Adults With Suspected Infection Admitted to the Intensive Care Unit. JAMA. 2017;317:290.

16. Kamaleswaran R, Lian J, Lin D-L, Molakapuri H, Nunna S, Shah P, et al. Predicting Volume Responsiveness Among Sepsis Patients Using Clinical Data and Continuous Physiological Waveforms. AMIA 2021 Annual Symposium. 2021.

17. Reyna MA, Josef CS, Jeter R, Shashikumar SP, Westover MB, Nemati S, et al. Early Prediction of Sepsis From Clinical Data: The PhysioNet/Computing in Cardiology Challenge 2019. Critical Care Medicine. 2020;48:210–7.

18. Moor M, Bennett N, Plečko D, Horn M, Rieck B, Meinshausen N, et al. Predicting sepsis using deep learning across international sites: a retrospective development and validation study. eClinicalMedicine. 2023;62:102124.

19. Chawla LS, Bellomo R, Bihorac A, Goldstein SL, Siew ED, Bagshaw SM, et al. Acute kidney disease and renal recovery: consensus report of the Acute Disease Quality Initiative (ADQI) 16 Workgroup. Nat Rev Nephrol. 2017;13:241–57.

20. Peerapornratana S, Priyanka P, Wang S, Smith A, Singbartl K, Palevsky PM, et al. Sepsis-Associated Acute Kidney Disease. Kidney International Reports. 2020;5:839–50.

21. Semler MW, Self WH, Wanderer JP, Ehrenfeld JM, Wang L, Byrne DW, et al. Balanced Crystalloids versus Saline in Critically Ill Adults. N Engl J Med. 2018;378:829–39.

22. Semler MW, Rice TW, Shaw AD, Siew ED, Self WH, Kumar AB, et al. Identification of Major Adverse Kidney Events Within the Electronic Health Record. J Med Syst. 2016;40:167.

23. Buuren SV, Groothuis-Oudshoorn K. mice: Multivariate Imputation by Chained Equations in R. J Stat Soft [Internet]. 2011 [cited 2025 Jan 2];45. Available from: http://www.jstatsoft.org/v45/i03/

24. Lee C, Yoon J, Schaar MVD. Dynamic-DeepHit: A Deep Learning Approach for Dynamic Survival Analysis With Competing Risks Based on Longitudinal Data. IEEE Trans Biomed Eng. 2020;67:122–33.

25. Vagliano I, Chesnaye NC, Leopold JH, Jager KJ, Abu-Hanna A, Schut MC. Machine learning models for predicting acute kidney injury: a systematic review and critical appraisal. Clinical Kidney Journal. 2022;15:2266–80.

26. Zarbock A, Küllmar M, Ostermann M, Lucchese G, Baig K, Cennamo A, et al. Prevention of Cardiac Surgery–Associated Acute Kidney Injury by Implementing the KDIGO Guidelines in High-Risk Patients Identified by Biomarkers: The PrevAKI-Multicenter Randomized Controlled Trial. Anesthesia & Analgesia [Internet]. 2021 [cited 2025 Jan 4]; Available from: https://journals.lww.com/10.1213/ANE.0000000000005458

27. Küllmar M, Weiß R, Ostermann M, Campos S, Grau Novellas N, Thomson G, et al. A Multinational Observational Study Exploring Adherence With the Kidney Disease: Improving Global Outcomes Recommendations for Prevention of Acute Kidney Injury After Cardiac Surgery. Anesthesia & Analgesia. 2020;130:910–6.

28. Kolhe NV, Staples D, Reilly T, Merrison D, Mcintyre CW, Fluck RJ, et al. Impact of Compliance with a Care Bundle on Acute Kidney Injury Outcomes: A Prospective Observational Study. James LR, editor. PLoS ONE. 2015;10:e0132279.

29. Justice CM, Nevin C, Neely RL, Dilcher B, Kovacic-Scherrer N, Carter-Templeton H, et al. Effect of Tiered Implementation of Clinical Decision Support System for Acute Kidney Injury and Nephrotoxin Exposure in Cardiac Surgery Patients. Appl Clin Inform. 2025;16:001–10.

30. Yu X, Xin Q, Hao Y, Zhang J, Ma T. An early warning model for predicting major adverse kidney events within 30 days in sepsis patients. Front Med. 2024;10:1327036.

31. Hochreiter S, Schmidhuber J. Long Short-Term Memory. Neural Computation. 1997;9:1735–80.

32. Yu Y, Si X, Hu C, Zhang J. A Review of Recurrent Neural Networks: LSTM Cells and Network Architectures. Neural Computation. 2019;31:1235–70.

33. Lee JM, Hauskrecht M. Recent Context-Aware LSTM for Clinical Event Time-Series Prediction. In: Riaño D, Wilk S, Ten Teije A, editors. Artificial Intelligence in Medicine [Internet]. Cham: Springer International Publishing; 2019 [cited 2025 Jan 3]. p. 13–23. Available from: http://link.springer.com/10.1007/978-3-030-21642-9_3

