## Supplementary Figure 1 and Table 1 for "ORAKLE: Optimal Risk prediction for mAke30 in patients with acute Kidney injury using deep Learning"

**SUPPLEMENTARY DOCUMENTS**

**Supplementary Figure S1.** Cohort inclusion and exclusion criteria.

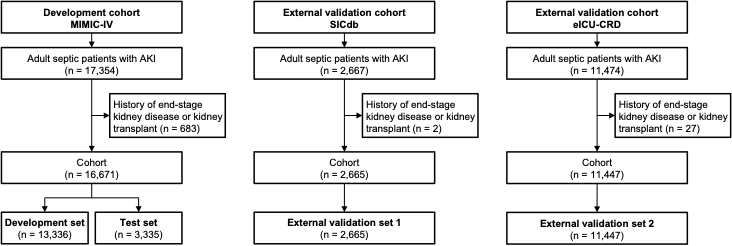

**Supplementary Table S1.** Detailed baseline characteristics of patients in the training and testing sets (MIMIC-IV) and external validation cohorts (SICdb and eICU-CRD).

|  | MIMIC-IV: Training set (N=13336) | MIMIC-IV: Test set (N=3335) | SICdb (N=2665) | eICU-CRD (N=11447) | Total (N=30783) | p value |
| --- | --- | --- | --- | --- | --- | --- |
| Heart rate, (mean±sd) | 87.12 ± 19.67 | 87.10 ± 19.39 | 83.91 ± 18.77 | 88.87 ± 20.39 | 87.49 ± 19.88 | < 0.001 |
| SBP, mmHg, (mean±SD | 112.98 ± 20.59 | 113.22 ± 20.76 | 107.42 ± 21.13 | 115.34 ± 23.98 | 113.40 ± 22.08 | < 0.001 |
| DBP, mmHg, (mean±SD | 60.21 ± 14.51 | 60.16 ± 14.28 | 54.29 ± 11.14 | 60.59 ± 15.39 | 59.84 ± 14.67 | < 0.001 |
| MBP, mmHg, (mean±SD | 75.14 ± 15.14 | 75.07 ± 14.92 | 72.41 ± 14.62 | 77.20 ± 18.12 | 75.67 ± 16.31 | < 0.001 |
| Respiratory rate, (mean±SD | 20.13 ± 5.72 | 19.96 ± 5.69 | 16.30 ± 5.55 | 20.65 ± 6.34 | 19.97 ± 6.05 | < 0.001 |
| Temperature, C, (mean±SD | 36.88 ± 0.76 | 36.90 ± 0.77 | 36.58 ± 1.91 | 36.83 ± 0.95 | 36.84 ± 0.99 | < 0.001 |
| SpO2, %, (mean±SD) | 96.67 ± 3.56 | 96.73 ± 3.42 | 96.15 ± 3.64 | 96.55 ± 4.17 | 96.58 ± 3.79 | < 0.001 |
| Weight, Kg, (mean±SD) | 84.55 ± 23.88 | 83.51 ± 23.41 | 80.94 ± 22.63 | 81.97 ± 20.35 | 83.17 ± 22.51 | < 0.001 |
| Hematocrit, %, (mean±SD) | 31.76 ± 6.25 | 31.74 ± 6.27 | 31.45 ± 6.37 | 32.89 ± 7.00 | 32.15 ± 6.58 | < 0.001 |
| Hemoglobin, g/dL, (mean±SD) | 10.41 ± 2.10 | 10.43 ± 2.11 | 10.66 ± 2.14 | 10.74 ± 2.33 | 10.56 ± 2.20 | < 0.001 |
| MCH, pg, (mean±SD) | 30.02 ± 2.80 | 30.07 ± 2.70 | 30.09 ± 2.46 | 29.82 ± 2.77 | 29.96 ± 2.75 | < 0.001 |
| MCHC, g/L, (mean±SD) | 32.78 ± 1.78 | 32.86 ± 1.74 | 33.93 ± 1.49 | 32.64 ± 1.60 | 32.84 ± 1.72 | < 0.001 |
| MCV, fL, (mean±SD) | 91.66 ± 7.52 | 91.59 ± 7.35 | 88.70 ± 6.27 | 91.35 ± 7.47 | 91.28 ± 7.43 | < 0.001 |
| Platelet, K/uL, (mean±SD) | 195.75 ± 111.33 | 192.24 ± 106.62 | 213.83 ± 106.40 | 192.83 ± 106.54 | 195.85 ± 108.78 | < 0.001 |
| RBC count, Count m/uL, (mean±SD) | 3.49 ± 0.73 | 3.49 ± 0.75 | 3.56 ± 0.73 | 3.62 ± 0.80 | 3.54 ± 0.76 | < 0.001 |
| WBC count, K/uL, (mean±SD) | 13.69 ± 11.42 | 13.35 ± 11.15 | 12.80 ± 7.16 | 13.86 ± 8.47 | 13.64 ± 10.06 | < 0.001 |
| Bicarbonate, mEq/L, (mean±SD) | 22.31 ± 5.19 | 22.36 ± 5.07 | 24.34 ± 4.61 | 22.69 ± 5.86 | 22.63 ± 5.42 | < 0.001 |
| BUN, mg/dL, (mean±SD) | 32.19 ± 25.23 | 31.65 ± 24.93 | 26.52 ± 21.05 | 36.79 ± 27.06 | 33.35 ± 25.75 | < 0.001 |
| Calcium, mg/dL, (mean±SD) | 8.24 ± 0.87 | 8.25 ± 0.93 | 8.47 ± 0.65 | 8.15 ± 0.93 | 8.23 ± 0.89 | < 0.001 |
| Serum creatinine, mg/dL, (mean±SD) | 1.74 ± 1.71 | 1.72 ± 1.72 | 1.51 ± 1.45 | 2.30 ± 2.19 | 1.92 ± 1.91 | < 0.001 |
| Sodium, mEq/L, (mean±SD) | 138.28 ± 5.64 | 138.45 ± 5.66 | 139.86 ± 5.06 | 138.61 ± 6.06 | 138.56 ± 5.77 | < 0.001 |
| INR, (mean±SD) | 1.56 ± 0.91 | 1.56 ± 0.88 | 1.33 ± 0.36 | 1.59 ± 1.07 | 1.55 ± 0.94 | < 0.001 |
| Total bilirubin, mg/dL, (mean±SD) | 1.90 ± 4.14 | 1.84 ± 4.33 | 1.21 ± 1.92 | 1.32 ± 2.63 | 1.62 ± 3.52 | < 0.001 |
| PO2, mm Hg, (mean±SD) | 108.17 ± 68.69 | 109.15 ± 71.39 | 94.68 ± 31.02 | 122.17 ± 80.61 | 112.31 ± 71.88 | < 0.001 |
| PCO2, mm Hg, (mean±SD) | 41.98 ± 11.84 | 41.71 ± 11.18 | 40.88 ± 9.67 | 40.83 ± 12.73 | 41.43 ± 11.95 | < 0.001 |
| Ph, (mean±SD) | 7.36 ± 0.09 | 7.36 ± 0.09 | 7.39 ± 0.08 | 7.34 ± 0.11 | 7.35 ± 0.09 | < 0.001 |
| Base excess, mEq/L, (mean±SD) | -1.87 ± 5.28 | -1.71 ± 5.17 | -0.59 ± 4.77 | -3.27 ± 7.06 | -2.26 ± 6.01 | < 0.001 |
| Vasopressor: Y/N | 4394 (32.9%) | 1079 (32.4%) | 1516 (56.9%) | 1426 (12.5%) | 8415 (27.3%) | < 0.001 |
| Vasopressor: Dose, mcg, (mean±SD) | 0.29 ± 0.95 | 0.30 ± 0.91 | 0.45 ± 0.93 | 0.93 ± 34.38 | 0.54 ± 20.98 | 0.094 |
| Mechanical ventilation: Y/N | 2925 (21.9%) | 747 (22.4%) | 466 (17.5%) | 3374 (29.5%) | 7512 (24.4%) | < 0.001 |
| Mechanical ventilation, Hour, (mean±SD) | 0.98 ± 2.18 | 1.01 ± 2.18 | 0.85 ± 2.08 | 1.09 ± 2.25 | 1.02 ± 2.20 | < 0.001 |
| Nephrotoxic drugs: Y/N | 1733 (13.0%) | 412 (12.4%) | 800 (30.0%) | 3186 (27.8%) | 6131 (19.9%) | < 0.001 |
